# Classification of Parkinson’s disease and essential tremor based on gait and balance characteristics from wearable motion sensors: A data-driven approach

**DOI:** 10.1101/2020.04.17.20065441

**Authors:** Sanghee Moon, Hyun-Je Song, Vibhash D. Sharma, Kelly E. Lyons, Rajesh Pahwa, Abiodun E. Akinwuntan, Hannes Devos

## Abstract

Parkinson’s disease (PD) and essential tremor (ET) are movement disorders that can have similar clinical characteristics including tremor and gait difficulty. These disorders can be misdiagnosed leading to delay in appropriate treatment. The aim of the study was to determine whether gait and balance variables obtained with wearable sensors can be utilized to differentiate between PD and ET using machine learning techniques. Additionally, we compared classification performances of several machine learning models. A balance and gait data set collected from 567 people with PD or ET was investigated. Performance of several machine learning techniques including neural networks (NN), support vector machine (SVM), k-nearest neighbor (kNN), decision tree (DT), random forest (RF), and gradient boosting (GB), were compared using F1-scores. Machine learning models classified PD and ET based on balance and gait characteristics better than chance or logistic regression. The highest F1-score was 0.61 of NN, followed by 0.59 of GB, 0.56 of RF, 0.55 of SVM, 0.53 of DT, and 0.49 of kNN. The results demonstrated the utility of machine learning models to classify different movement disorders. Further study will provide a more accurate clinical tool to help clinical decision-making.

## 1. Introduction

Parkinson’s disease (PD) and essential tremor (ET) are common movement disorders characterized by the presence of tremor [1]. Although ET has traditionally been considered a mono-symptomatic disorder presenting with tremor, increasing evidence suggests that ET is a complex disorder with involvement of other motor and non-motor symptoms [2]. Both PD and ET can share clinical features including motor symptoms such as bradykinesia (slow movement), gait impairment and dystonia (involuntary muscle contraction), and non-motor symptoms such as cognitive impairments, sleep disturbances, depression, and anxiety [3, 4]. Diagnosis of these disorders can be challenging for clinicians due to overlapping symptoms, and these disorders are frequently confused and misdiagnosed. A past study reported that about a third of patients with PD or dystonia were misdiagnosed with ET [5]. Since misdiagnosis can prevent or delay appropriate medical care and worsen patients’ quality of life, accurate differentiation between PD and ET is important to provide optimal care.

Clinical observation of gait and balance impairments can play a major role in classifying different conditions and monitoring the progression of PD and ET. Subtle changes in gait have even been found to occur before a clinical diagnosis of PD [6, 7], Alzheimer’s disease [8], or multiple sclerosis [9], suggesting gait as a potential biomarker for neurological disorders. Balance and gait impairments are more prominent and clinically observable in PD than in ET. However, there is growing evidence suggesting gait abnormalities in patients with ET [10]. Previous studies showed gait and balance abnormalities such as decreased cadence [11, 12], decreased gait speed [12], increased double support [11, 12], abnormalities in tandem gait [13-15], and postural instability in ET [11, 16]. These abnormalities in ET are also commonly found in PD, which contribute to misdiagnosis of the two movement disorders [5].

Advances in technology enable an objective assessment of gait and balance through numerous devices such as body-worn inertial motion unit (IMU) sensors, 3-dimentional motion capturing systems, force plates, gait walkways, and smartphones. Many movement disorder clinics and research laboratories have started to implement these technological devices in their practices [17], particularly IMU sensors to evaluate balance and gait in PD [18, 19]. Subsequently, a vast amount of complex and non-linear data from technological devices are available for clinicians and researchers that require advanced statistical analyses. Machine learning is widely employed to analyze large data sets produced from movement disorder clinics and research laboratories [20]. Among various machine learning techniques, neural network (NN) models are broadly utilized due to their superior performance compared to traditional analytic methods such as logistic regression (LR) [21]. Previously, NNs have been employed in gait and balance studies to process signals from wearable devices in PD [22-25]. In addition, a study used a NN to successfully discern PD from ET using surface electromyography data [26]. Other machine learning algorithms such as support vector machine (SVM) and k-nearest neighbor (kNN) were used to differentiate between PD and ET based on IMU sensors, but they mainly investigated upper body tremors [27-30]. To our knowledge, no study has utilized machine learning techniques to differentiate between PD and ET based on data collected from gait and balance characteristics from wearable IMU sensors.

Therefore, the primary aim of this data-driven study was to distinguish between PD and ET using balance and gait characteristics obtained from IMU sensors using machine learning techniques. The secondary aim of the study was to compare and evaluate different machine learning performances in distinguishing between PD and ET using balance and gait data obtained from IMU sensors.

## 2. Methods

### 2.1. Participants

The instrumented gait and balance tests were administered at the Parkinson’s Disease and Movement Disorder Clinic of the University of Kansas Medical Center. Between January 3, 2017 and December 11, 2018, a total of 1,468 people was tested. We excluded people if they were diagnosed with both PD and ET (n = 29) and/or if they had a history of deep brain stimulation surgery (n = 468). For those who visited the clinic more than once during the study period (n = 628), we only included the data from their first visits. Additionally, we excluded people with no data recorded due to technical error of the measuring device (n = 65), leaving a total of 567 people with PD or ET in the study.

### 2.2. Protocol and materials

Participants wore six IMU sensors (Opal, APDM, Inc., Portland, OR, USA). Two wrist sensors were bilaterally mounted on the dorsal side of the wrist and two foot sensors were bilaterally mounted to the instep (dorsal side of metatarsus) of each foot. The sternum sensor was mounted on the sternum of the chest and the lumber sensor was mounted to the posterior side at the L5 region. All six sensors were firmly tightened to the designated locations using straps during testing.

The instrumented stand and walk (iSAW) test was administered. During the iSAW test, participants were instructed to stand still for 30 seconds, walk straight for 7 meters at a comfortable speed after hearing a beep, turn 180° around at the end of 7-meter marker, then walk back to the start point. The iSAW test is a reliable and valid balance and gait measure for clinical use [31-33].

The IMU utilized in the study contained two accelerometers (range: ± 16g and ± 200g, resolution: 14 and 17.5 bits, sample rate: 128 Hz), a gyroscope (range: ± 2000°/s, resolution: 16 bits, sample rate: 128 Hz), and a magnetometer (range: ± 8 Gauss, resolution: 12 bits, sample rate: 128 Hz). A total of 130 gait and balance parameters were automatically computed using the Mobility Lab software (APDM, Inc., Portland, OR, USA).

### 2.3. Data analysis

#### 2.3.1. Pre-processing data set

Among 130 gait and balance parameters computed by the Mobility Lab software, a total of 48 parameters with clinical relevance were included in the study based on a recent review [34] and the clinical expertise of the authors (Table 1). In the data set, 524 participants had PD (age = 66.73 ± 9.17, disease duration = 8.20 ± 5.11 years), while 43 had ET (age = 66.98 ± 9.84, disease duration = 13.83 ± 13.79 years) as their primary diagnoses. The ratio between PD participants and ET participants was highly imbalanced, 92.5% (PD) vs 7.5% (ET), in the data set. To mitigate the effect derived from the imbalanced data, we utilized a Synthetic Minority Over-sampling Technique (SMOTE) [35], an oversampling approach to create synthetic minority class examples. For missing values, we used a univariate feature imputation algorithm to predict missing values in datasets before training the classification model. We initially attempted to reduce the dimension of the data using principal component analysis (PCA); however, we found that there was a slight performance drop when applying PCA. One potential reason behind this phenomenon was that we manually selected useful features before PCA. Hence, we did not utilize the dimension reduction technique.

**Table 1.** Gait and balance features utilized in the analysis.

|  |  |
| --- | --- |
| **Gait – lower limb** |  |
| Cadence [left] (steps/min) | Cadence [right] (steps/min) |
| Double support [left] (%GC) | Double support [right] (%GC) |
| Gait speed [left] (m/s) | Gait speed [right] (m/s) |
| Lateral step variability [left] (cm) | Lateral step variability [right] (cm) |
| Foot strike angle [left] (degrees) | Foot strike angle [right] (degrees) |
| Toe off angle [left] (degrees) | Toe off angle [right] (degrees) |
| Single limb support [left] (%GC) | Single limb support [right] (%GC) |
| Stance [left] (%GC) | Stance [right] (%GC) |
| Step duration [left] (s) | Step duration [right] (s) |
| Stride length [left] (m) | Stride length [right] (m) |
| Swing [left] (%GC) | Swing [right] (%GC) |
| Terminal double support [left] (%GC) | Terminal double support [right] (%GC) |
| **Gait – lumbar** |  |
| Coronal range of motion (degrees) |  |
| Sagittal range of motion (degrees) |  |
| Transverse range of motion (degrees) |  |
| **Gait – trunk** |  |
| Coronal range of motion (degrees) |  |
| Sagittal range of motion (degrees) |  |
| Transverse range of motion (degrees) |  |
| **Postural sway** |  |
| Mean velocity (m/s) | Acc - path length (m/s<sup>2</sup>) |
| Mean velocity [coronal] (m/s) | Acc - path length [coronal] (m/s<sup>2</sup>) |
| Mean velocity [sagittal] (m/s) | Acc - path length [sagittal] (m/s<sup>2</sup>) |
| Acc - RMS sway (m/s<sup>2</sup>) | Acc - RMS sway (degrees) |
| Acc - RMS sway [coronal] (m/s<sup>2</sup>) | Acc - RMS sway [coronal] (degrees) |
| Acc - RMS Sway [sagittal] (m/s<sup>2</sup>) | Acc - RMS sway [sagittal] (degrees) |
| Sway area radius [coronal] (degrees) | Acc - range (m/s<sup>2</sup>) |
| Sway area rotation (degrees) | Acc - range [coronal] (m/s<sup>2</sup>) |
| Sway area (degrees<sup>2</sup>) | Acc - range [sagittal] (m/s<sup>2</sup>) |

Abbreviation: Acc = acceleration, GC = gait cycle, RMS = root mean square

#### 2.3.2. Classification and model selection

The classification models included NN, SVM, kNN, decision tree (DT), random forest (RF), gradient boosting classifier (GB), and LR. To find out the optimal values of the hyper-parameters of the classification models, we used a stratified 3-fold cross validation with grid search strategy. Table 2 shows the hyper-parameter search spaces of each classification model.

**Table 2.** Model hyper-parameters of the classification models.

| Classification models | Hyper-parameter search spaces |
| --- | --- |
| Neural network (NN) | hidden_layer_sizes = {100, 200, 300} |
| Support vector machines (SVM) | C = {0.01, 0.1, 1, 5, 10, 100}, kernel = {'linear', 'rbf'},<br>gamma = {0.01, 0.1, 1, 10}, class_weight = {None, 'balanced'} |
| k-nearest neighbor (kNN) | n_neighbors = {1,3,5,7,9}, weights = {'uniform', 'distance'} |
| Decision tree (DT) | max_depth = {5, 6, 7, 8, 9, 10, 15, 20}, class_weight = {None, 'balanced'} |
| Random forest (RF) | n_estimators = {20, 50, 100, 200}, class_weight = {None, 'balanced', 'balanced_subsample'} |
| Gradient boosting (GB) | n_estimators = {20, 50, 100, 200} |
| Logistic regression (LR) | C = {0.01, 0.1, 1, 5, 10, 100}, penalty = {'l1', 'l2'},<br>class_weight = {None, 'balanced'} |

Note: gamma parameter in SVM was applied when the kernel is radial basis function 'rbf'; class\_weight was applied when the oversampling approach (SMOTE) was not used.

#### 2.3.3. Performance evaluation

The classification models were evaluated with accuracy (a ratio of correct prediction to total observations), recall (a ratio of correct prediction of positive cases to all observations in actual cases), precision (a ratio of correct prediction of positive cases to all positive cases), and F1 score (a harmonic mean of precision and recall). Of note, all performances were micro-averaged. The accuracy, and precision and recall for the F1-score were calculated as follows (TP = true positive, TN = true negative, FP = false positive, FN = false negative):

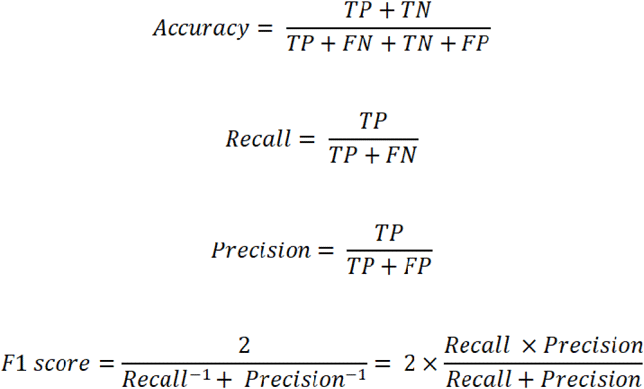

## 3. Results

The results of NN, SVM, kNN, DT, RF, GB, and LR with and without the oversampling approach are shown in Table 3. With SMOTE, (1) the accuracy of the models ranged from 0.65 (kNN) to 0.89 (NN); (2) the precision was similar across the models ranging from 0.54 (SVM, kNN, DT, and LR) to 0.61 (NN); (3) the recall ranged from 0.58 (DT) to 0.63 (kNN and GB); and (4) the F1-score ranged from 0.53 (DT and LR) to 0.61 (NN).

**Table 3.**
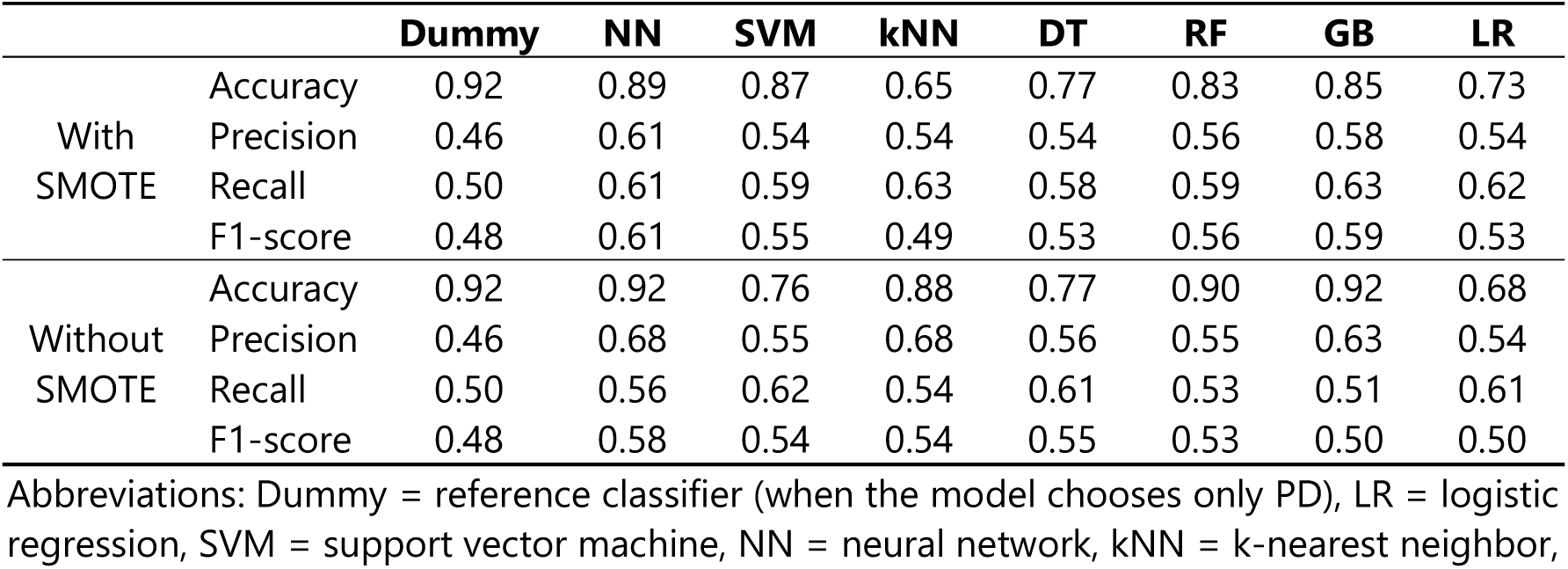
Accuracy, Precision, Recall, and F1-score of logistic regression, support vector machine, neural network, k-nearest neighbor, decision tree, random forest, and gradient boosting.

|  |  | Dummy | NN | SVM | kNN | DT | RF | GB | LR |
| --- | --- | --- | --- | --- | --- | --- | --- | --- | --- |
| With SMOTE | Accuracy | 0.92 | 0.89 | 0.87 | 0.65 | 0.77 | 0.83 | 0.85 | 0.73 |
|  | Precision | 0.46 | 0.61 | 0.54 | 0.54 | 0.54 | 0.56 | 0.58 | 0.54 |
|  | Recall | 0.50 | 0.61 | 0.59 | 0.63 | 0.58 | 0.59 | 0.63 | 0.62 |
|  | F1-score | 0.48 | 0.61 | 0.55 | 0.49 | 0.53 | 0.56 | 0.59 | 0.53 |
| Without SMOTE | Accuracy | 0.92 | 0.92 | 0.76 | 0.88 | 0.77 | 0.90 | 0.92 | 0.68 |
|  | Precision | 0.46 | 0.68 | 0.55 | 0.68 | 0.56 | 0.55 | 0.63 | 0.54 |
|  | Recall | 0.50 | 0.56 | 0.62 | 0.54 | 0.61 | 0.53 | 0.51 | 0.61 |
|  | F1-score | 0.48 | 0.58 | 0.54 | 0.54 | 0.55 | 0.53 | 0.50 | 0.50 |

Abbreviations: Dummy = reference classifier (when the model chooses only PD), LR = logistic regression, SVM = support vector machine, NN = neural network, kNN = k-nearest neighbor,
RF = random forest, GB = gradient boosting, SMOTE = synthetic minority over-sampling technique.

## 4. Discussion

This data-driven study aimed to differentiate between two movement disorders, PD and ET based on gait and balance characteristics collected from IMU sensors using various machine learning models. Additionally, the classification performance was compared across different machine learning models.

Recent technological advances enable clinics and research laboratories to employ wearable devices in their balance and gait assessments. This allows precise measurement of balance and abnormalities and accurate monitoring of physical activities of daily living. However, the data produced by technological devices are often overwhelming and under-utilized due to the size and complexity of the data [36]. Our data set provides useful clinical information for gait and balance such as gait speed, cadence, and postural sway collected by wearable devices. Our current results suggest that machine learning models can increase the utility of these complex data collected by technological devices. Our machine learning models outperformed (F1-scores ranging between 0.49 and 0.61) the dummy model (F1-score = 0.48) to classify the two movement disorders.

In this study, with SMOTE, the NN outperformed other models in classifying two different movement disorders solely based on a clinically available gait and balance data set. The F1-score of NN was 0.61, showing the highest performance among 8 models in the analysis. The robustness of NN performance typically shows in large and complex data. Previous studies in PD have demonstrated NN as the superior machine learning technique using data collected from wearable IMU sensors in levodopa-induced dyskinesia assessment and detection [22, 23], gait abnormality classification [24], and discrimination between people with PD who underwent subthalamic stimulation and healthy controls [25]. Although the GB showed a similar performance based on the F1-score (0.59), the accuracy of GB (85%) was lower than that of the NN (89%). Accuracies of other comparison models including SVM, kNN, DT, RF, and LR ranged between 65% and 87% and their F1-scores were lower than the NN. However, particularly for this data-driven study, a higher performance in accuracy is not likely to reflect superior performance of the model, because the data set was heavily imbalanced with 92.5% of people with PD and 7.5% of people with ET. This implies that a dummy model will be 92.5% accurate in classifying PD if the model categorized each case as PD. The F1-score is a more adequate measure especially for an imbalanced data set in machine learning, because a greater F1-score reflects both low false positives and negatives [37].

The current study has several limitations. First, in our study, the data set was imbalanced towards an overrepresentation of PD. To overcome this limitation, we implemented the SMOTE that generates synthetic minority class samples, which is a widely used oversampling method [38]. Our findings demonstrated the SMOTE increased the classification performance, based on F1-scores, in the majority of models in the study (Table 3). This result may indicate the SMOTE was effective to minimize the influence of imbalanced class distribution in the current data set. However, further classification studies are required to use a data set that better reflects the actual disease representation in the population. Second, the design of the current study was a cross-sectional design including patients with clinically diagnosed PD or ET. Future research needs to include a longitudinal analysis using data over time to inform the accuracy of NNs using gait and balance characteristics from wearable devices to assist in the differential diagnosis or disease progression of PD and ET. Third, unlike past studies that utilized raw signal data captured by IMU sensors [22-25], the current study utilized pre-processed data (i.e. gait speed, sway area, cadence, etc.) from raw signal data as input variables. We opted to use the pre-processed data since these are readily available, adding to the clinical relevance of our current findings. However, we acknowledge that using raw data adds more information to the model since the pre-processing procedure might result in a significant loss of raw signal features directly from IMU sensors. In general, the performance of NN can be more precise and accurate when the model is fed more data. Thus, further examination using raw data collected from wearable IMU sensors can offer new insights that extend our current findings.

## 5. Conclusions

Wearable sensors for balance and gait assessments can be implemented in movement disorders clinics to produce a vast amount of potentially informative data for assisting in diagnosis and monitoring disease progression. The current study showed that NN with SMOTE outperformed machine learning models and traditional logistic regression in classifying PD and ET based on the pre-processed balance and gait IMU data set. With further validation, a data-driven approach using machine learning techniques may provide a more efficient diagnostic and prognostic tool that can aid clinicians’ decision-making process.

## Data Availability

Data will be made available on request.

## Ethical Statement

The study ethics was approved by the University of Kansas Medical Center Institutional Review Board (#12351 - Movement Disorder Research Registry and #STUDY00143000 - Gait assessment in Movement Disorders). The data set used in this retrospective data base study was collected as part of routine patient care after patient’s consent.

## Author Contributions

Conceptualization, S.M., H.D., V.D.S., K.E.L. and R.P.; methodology, S.M., H-J.S. and H.D.; software, S.M. and H-J.S.; formal analysis, S.M. and H-J.S.; investigation, S.M., H-J.S., V.D.S., K.E.L, R.P., A.E.A and H.D.; resources, V.D.S., K.E.L. and R.P.; data curation, S.M. and H-J.S.; writing—original draft preparation, S.M.; writing—review and editing, H-J.S., H.D., A.E.A, V.D.S., K.E.L. and R.P.; supervision, H.D.; project administration, S.M.; funding acquisition, V.D.S. All authors have read and agreed to the published version of the manuscript.

## Funding

This research received grant from the International Essential Tremor Foundation for Gait Assessment in Essential Tremor Using Wireless Sensors (V.D.S).

## Conflicts of Interest

The authors declare no conflict of interest.

## Notes

### Competing Interest Statement

The authors have declared no competing interest.

